# Fall 2021 Resurgence and COVID-19 Seroprevalence in Canada Modelling waning and boosting COVID-19 immunity in Canada A Canadian Immunization Research Network Study

**DOI:** 10.1101/2021.08.17.21262188

**Authors:** David W. Dick, Lauren Childs, Zhilan Feng, Jing Li, Gergely Röst, David L Buckeridge, Nick H Ogden, Jane M Heffernan

## Abstract

There is a threat of COVID-19 resurgence in Fall 2021 in Canada. To understand the probability and severity of this threat, quantification of the level of immunity/protection of the population is required. We use an age-structured model including infection, vaccination and waning immunity to estimate the distribution of immunity to COVID-19 in the Canadian population. By late Summer 2021, coinciding with the end of the vaccination program, we estimate that 60 *−* 80% of the Canadian population will have some immunity to COVID-19. Model results show that this level of immunity is not sufficient to stave off a Fall 2021 resurgence. The timing and severity of a resurgence, however, varies in magnitude given multiple factors: relaxation of non-pharmaceutical interventions such as social distancing, the rate of waning immunity, the transmissibility of variants of concern, and the protective characteristics of the vaccines against infection and severe disease. To prevent large-scale resurgence, booster vaccination and/or re-introduction of public health mitigation may be needed.

## Introduction

The COVID-19 pandemic continues to affect the lives of Canadians. Despite increasing vaccination uptake, of first and second doses, and great decreases in COVID-19 cases of late, questions remain as to the future of the COVID-19 pandemic in this country. These centre on understanding the future of COVID-19 immunity, and possibilities of resurgence in the short and long term. i.e, can the Canadian population reach herd immunity?; is there risk of future resurgence in Fall 2021 - Winter 2022, or later?

A needed step in answering these questions lies in the determination and quantification of immunity in the Canadian population. Seroprevalence studies in different population cohorts have been conducted (see (1; 2) for examples, and (3) for more information), and many others are underway (see (4) for details). Such studies can be used to inform immunity distribution calculations. A recent statistical study by the COVID-19 Immunity Task Force (CITF), which incorporates different population seroprevalence studies into their analysis, estimated that the Canadian population has some immunity against COVID-19: due to infection, 5.4% (95% CrI: 0.6 to 15.8) as of May 31, 2021; due to infection and vaccination, 44.9% (95% CrI: 44.2 to 45.8) as of same date (5). The CITF (5) is the only working group that we know of that has attempted to quantify seroprevalence in the Canadian population.

It is of interest to provide other estimates of COVID-19 immunity in the Canadian population. Mathematical models of COVID-19 infection and vaccination can be used to estimate immunity distributions. In a previous study, we developed a mathematical model of COVID-19 infection and vaccination (6). The model tracks infection and immunity status by age. An important difference of our model over other models of COVID-19 is that it incorporates differential outcomes of immunity, determined by infection severity, which is ultimately related to the prevalence of comorbidities in the Canadian population by age. A second important difference is that our model includes the effects of waning immunity, whereby immunity protection can decay over time, which is a known characteristic of coronaviruses.

In the current study we use our mathematical model to determine distributions of immunity in the Canadian population, by age, from infection, and from vaccination. The model is fit to daily COVID-19 incidence data up to June 27, 2021(7), and incorporates actual (up to June 27, 2021) and projected (to September 2021) coverage of the first and second doses of COVID-19 vaccines (7; 8). We then use the model to quantify distributions of immunity from January 2020 to March 2022, given different assumed characteristics of the vaccines against various variants of concern (i.e., protection from infection, protection from severe disease), and different rates of waning immunity. In summary, we find that the magnitude of COVID-19 resurgence in Fall 2021 - Winter 2022 is sensitive to the rate of waning immunity, relaxation of non-pharmaceutical interventions and social distancing measures, the transmissibility of the virus, and on vaccine effectiveness against infection and disease.

## Methods

We implemented a model of COVID-19 infection with age structure (i.e., groups 0-4, 5-9, …, 75+ years). A flow diagram of the model is shown in Figure 1 for one age group. The model is based on a Susceptible-Exposed-Infected-Vaccinated-Susceptible model structure (SEIVS). We use 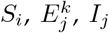, and 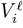 to denote the number of susceptible, exposed, infected and vaccinated individuals in each age group, where *i* (1 ≤ *i* ≤ 4) denotes immune status, *j* (2 ≤ *j* ≤ 4) denotes symptom severity, *k* (1 ≤ *k* ≤ 3) represents stages in the exposed class (to obtain Gamma-distributed exposed sojourns), and *l* = 1, 2 denotes the number of doses of vaccine that individuals have received.

**Figure 1:**
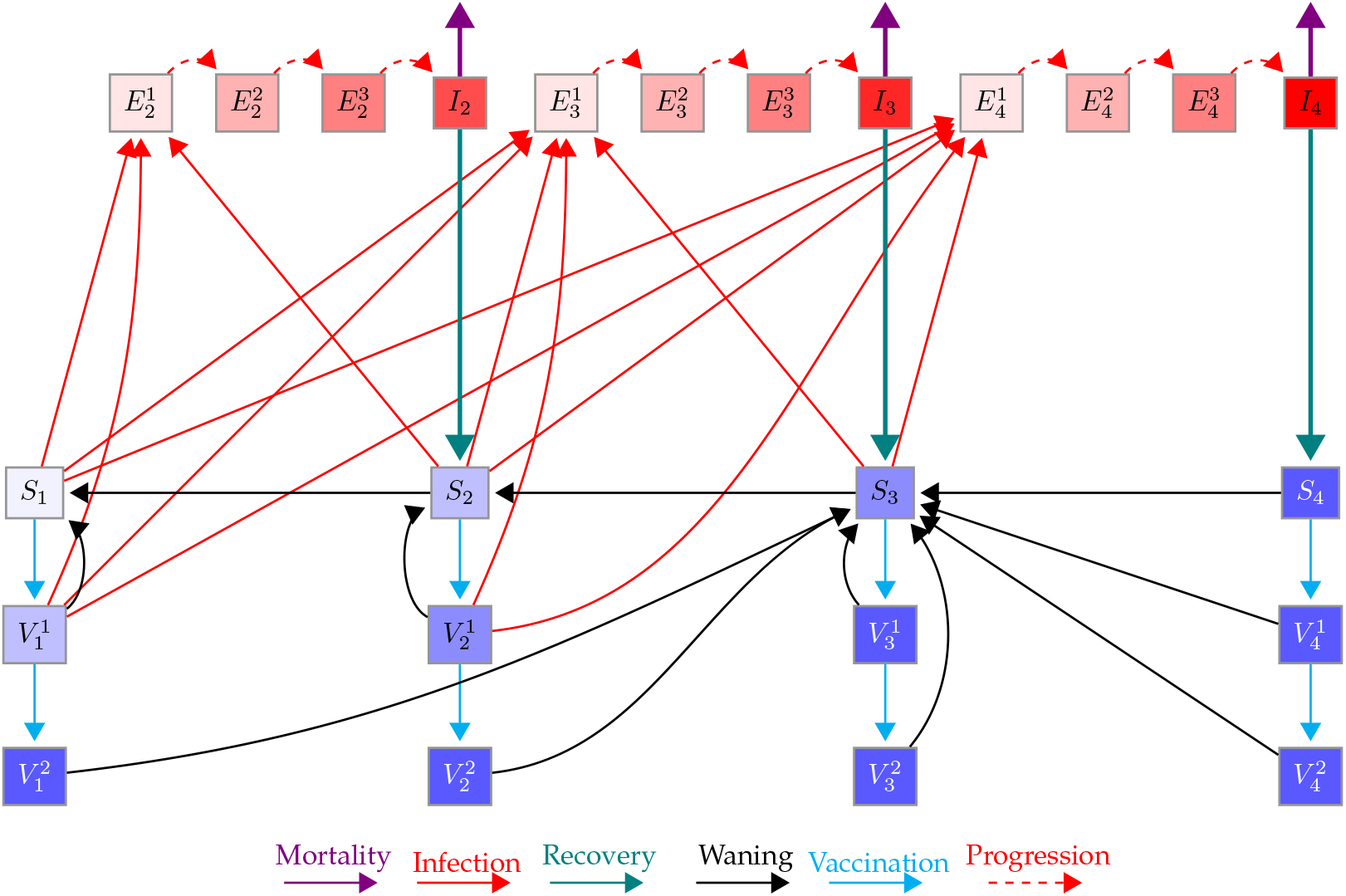
Schematic of the model for one age group. Here, *S*_1_, *S*_2_, *S*_3_, and *S*_4_ (purple shaded boxes) represent susceptible individuals who are immunologically naive, have some, moderate, and full immunity, respectively. *I*_2_, *I*_3_, and *I*_4_ (red boxes) represent infected individuals with mild, moderate and severe symptoms, respectively, who will develop some, moderate, and full immunity once recovered (teal solid line), respectively. 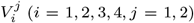 represent vaccinated individuals from the *S*_*i*_ classes (*i* = 1, 2, 3, 4) after *j* = 1, 2 doses of vaccine given a two-dose schedule. 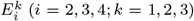 represent exposed individuals (infected, asymptomatic, not infectious) with progressive stages *k* = 2, 3, 4 that will experience mild *I*_2_, moderate *I*_3_, and severe *I*_4_ symptoms. Susceptible and vaccinated individuals can be infected and move to the exposed classes (red lines). Susceptible and vaccinated classes at the same location on the immunity continuum have similar characteristics. Immunity gained from infection and vaccination can wane (black lines).

A detailed description of the mathematical model can be found in (6). Briefly, we assume that mild (*I*_2_), moderate (*I*_3_), or severe (*I*_4_) disease can be experienced upon infection, and that the probability of mild, moderate, or severe disease is determined by the comorbidity status in each age group (9). We assume that all *I*_4_ infecteds will be reported. We also assume that some fraction of *I*_3_ will get tested and will be reported. Finally, we account for a small number of reported cases in *I*_3_ and *I*_2_ that will be tested because of contact tracing.

We assume that immunity gained after infection correlates with the severity of infection, such that higher levels of immunity are gained in individuals that have experienced more severe disease (10–12). Immunity gained from vaccination is also implemented in the model, using actual Canadian vaccination roll-out data and projections (7; 8). Three different types of vaccine characteristics are considered which reflect protective capacities against infection and/or disease (given different variants of concern (VOC)) of the vaccines used in the Canadian population (13–19). Characteristics of the vaccines are listed in Table 1.

**Table 1:** Vaccine efficacy. Three considered vaccine’s first and two-dose efficacy against infection and against severe disease.

|  | Against infection |  | Against severe disease |
| --- | --- | --- | --- |
|  | Two doses | First dose |  |
| Vaccine 1 | 70 % | 50 % | 75 % |
| Vaccine 2 | 80 % | 70 % | 80 % |
| Vaccine 3 | 90 % | 70 % | 92 % |

In addition to modelling the gain of immunity, we also consider immunity decay in the population. Different waning rates are considered such that immunity wanes on average 1 year or 3 years between *S* and *V* classes, and consecutive *S* classes, which gives waning from full immunity to full susceptibility over 3 years or 9 years, respectively. For comparison, we also consider the case when immunity does not wane.

Finally, public health mitigation is incorporated into the model using modified contact matrices for home, school, work, and other types of contacts between age groups. Figure 7 plots the mitigation windows (green vertical lines) and the percent reduction in contacts gained from the contact matrix mitigation modifications in each window (x’s). The model also collectively accounts for compliance to social distancing and mask wearing, changes in testing and contact tracing rates, and changes in transmission due to VOCs and weather. This is done using a parameter *κ* that is determined from a model fit to daily incidence data.

Using our mathematical model, we track the distribution of immunity in the Canadian population over time, given different characteristics of protective capacity from the vaccine, and different assumptions with respect to the waning rate of immunity. There are nine scenarios that we consider altogether. We fit the model for each scenario from January 25, 2020 to June 27, 2021.

We modify the contact matrices corresponding to phase 1 in June to represent easing of lockdown restrictions across Canada with the schools remaining closed. In July, we increase contacts in public spaces, to reflect different reopening steps taken in Canadian jurisdictions. In September the contact restrictions are eased to phase 2, representing school reopening and easing of restrictions at work. For a detailed description of the mitigation phases see Table 2.

Behaviour is likely to relax over the summer and into the fall, increasing transmission. In July 2021 many jurisdictions relaxed social distancing rules. To reflect this, and the prevalence of the delta VOC, we increase *κ* by a factor of 2. Reductions in transmissibility due to weather should only affect the summer months. We suggest, therefore, that *κ* can be increased in September 2021. We compare model results with no change in *κ* in September 2021, a 20% increase to reflect increased transmissibility due to changes in the weather (20), and a 38% increase (to allow for consideration of introduction of a new VOC - see Figure 7).

## Results

### Model Fit

Figures 2, 8, and 11 show the daily incidence of severe, moderate + severe, and mild + moderate + severe infections for the model fitting for all nine scenarios. Daily incidence data is shown in red, with the last day of the fitting denoted by the red vertical line. The remaining data points show appropriate model trend. We note that the model fit is the same in each of these figures. The difference between Figures 2, 8, and 11 lies only from September 2021, when school opens, and variable increases in *κ* are implemented. From September 2021 we consider three different scenarios for *κ*: no predicted change from the last estimated value (Figure 8), a 20% increase in *κ* (Figure 2), and a 38% increase (Figure 11).

**Figure 2:**
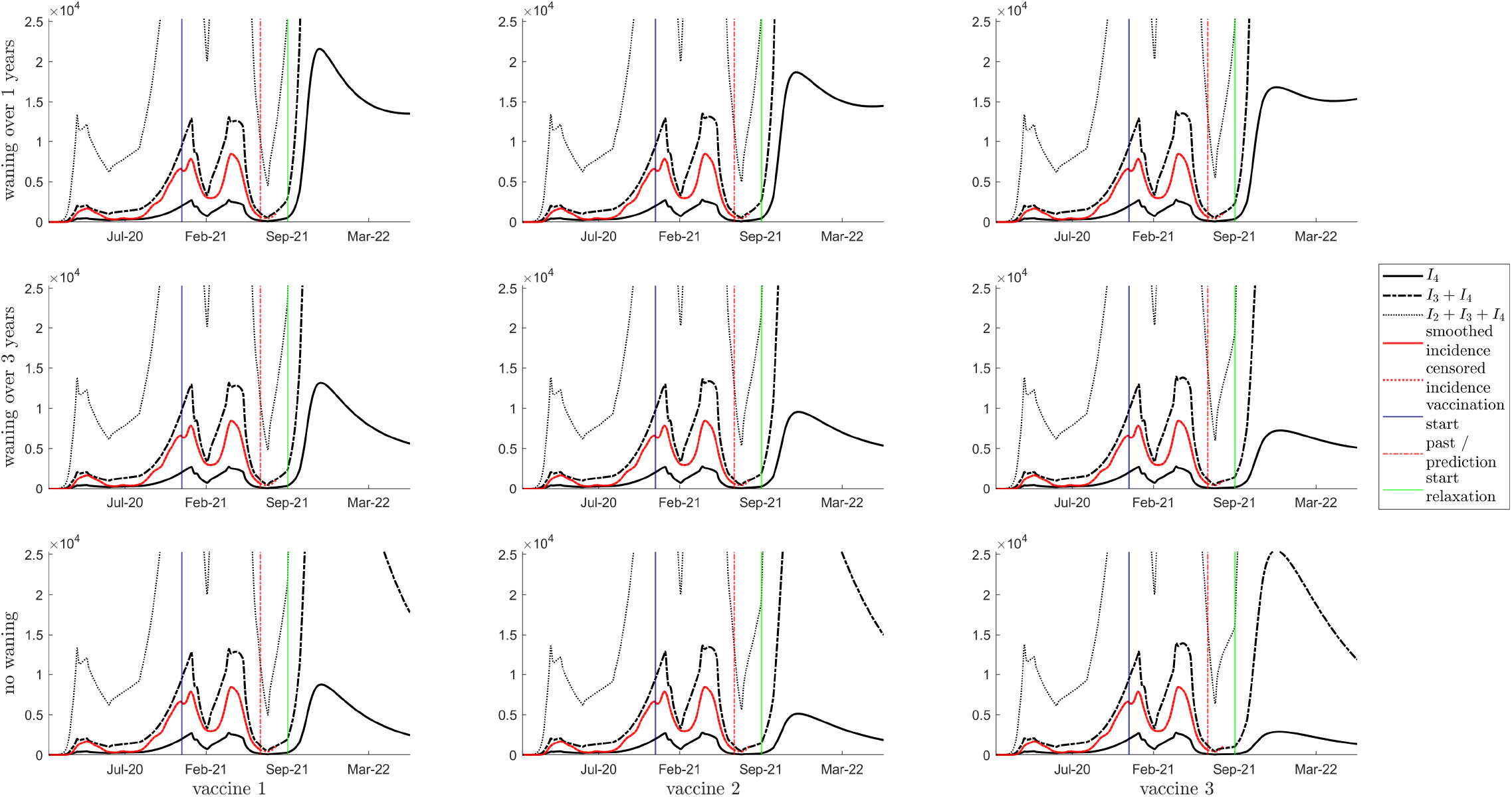
Simulation results with a 20% relaxation of *κ* starting in September 2021 showing daily incidence of *I*_4_ (solid line), *I*_3_ + *I*_4_ (dashed line) and *I*_2_ + *I*_3_ + *I*_4_ (dotted line), and smoothed incidence data is shown with a solid red line. The vertical blue, red and green lines indicate the start of vaccination, the beginning of the prediction phase, the start of relaxation, respectively. The top row is waning of immunity by one year between consecutive classes; the middle row is waning of immunity of three years; the bottom row is no waning of immunity. Columns left to right represent vaccines 1 to 3, respectively.

The fitted values of *κ* are shown in Figure 7 (bottom panel) considering each vaccination and waning scenario. In Figure 7 (top panel), we plot the percent reduction in contacts from contact matrix modifications and *κ* (red +’s) for Vaccine 1 with a waning rate *ω* = 1/year (see Table 1 and dashed blue line in Figure 7 bottom panel).

### Seroprevalence

Figures 3, 9 and 12 provide measurements of seroprevalence in the Canadian population given all vaccine types (Table 1) and changes in *κ* from September 2021 ((assuming *κ* values increase by 20%, no change, and 38%, respectively). These figures show all classes that confer some immunity to SARS-CoV-2, over all age groups. The colours denote the sum of the one-dose vaccinated sub-populations 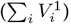, the two-dose vaccinated sub-populations 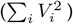, and the susceptible classes with partial and full immunity (*S*_2_ + *S*_3_ + *S*_4_), over 10-year age groups.The shading is related to the age classes, with lighter to darker coinciding with younger to older ages. In all nine scenarios, 60 to 80 % of the Canadian population will have some immunity against the pathogen (derived from infection and/or vaccination) by the time schools re-open in September 2021.

**Figure 3:**
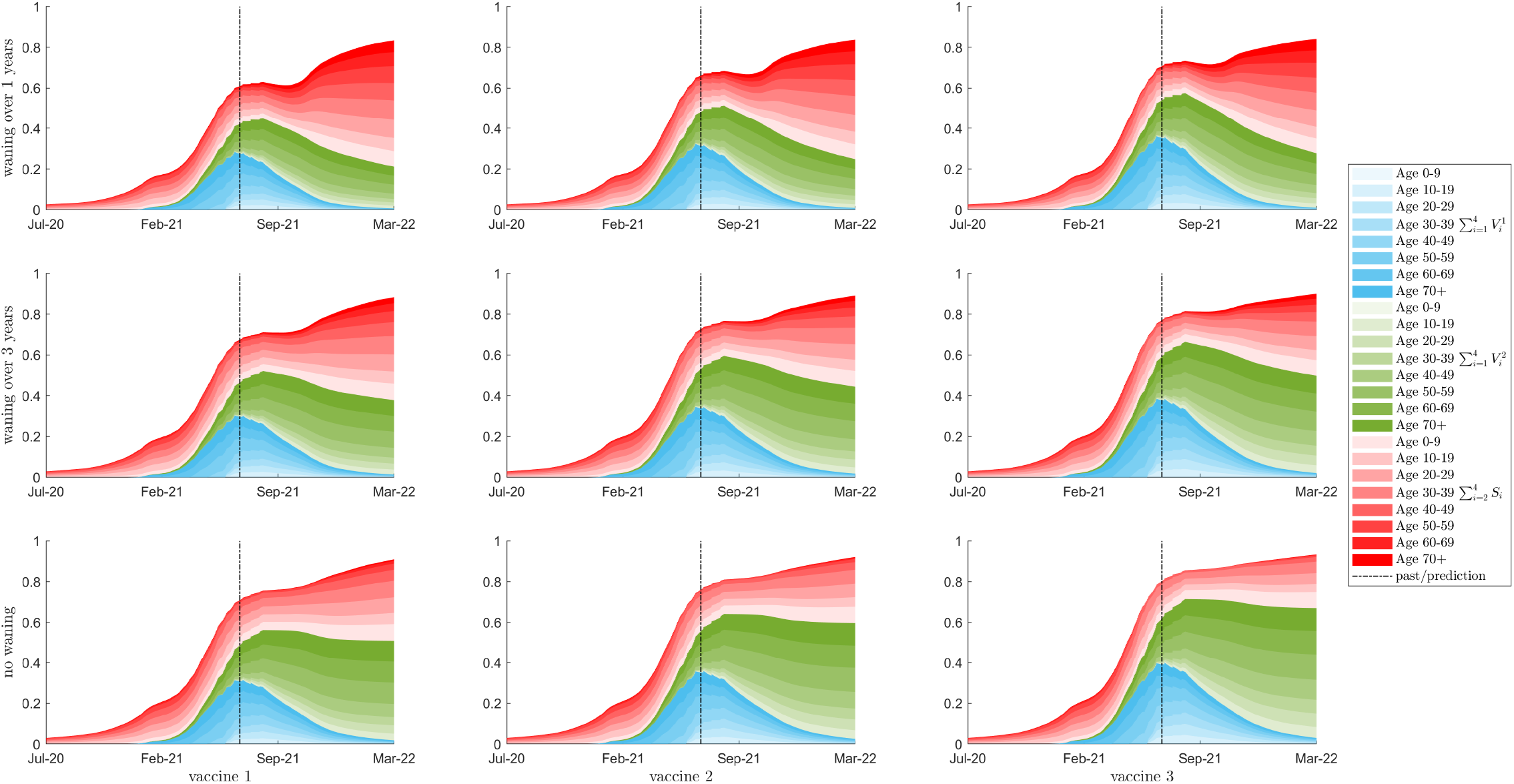
Simulation results with a 20% relaxation of *κ* starting in September 2021 showing seroprevalence as a percent of the total population for 10 year age classes are shown with colour intensity corresponding to age class. The red region is the sum of a susceptible classes that have been exposed to the virus, either from natural infection or through waning from the vaccinated classes. The blue and green regions show the populations of the first and second dose vaccinated classes respectively. The total population with some immunity (the top of the red region) is equal to the vertical sum of the three Blue, Red, and Green regions. The top row is waning of immunity by one year between consecutive classes; the middle row is waning of immunity of three years; the bottom row is no waning of immunity. Columns left to right represent vaccines 1 to 3, respectively. The vertical bar denotes the beginning of the prediction phase.

While the seroprevalence predicted by our model is higher than that projected by (5), there is little difference in the vaccine derived immunity between our results. The difference thus lies in the estimates of immunity derived from infections. While our model includes age structure, we must note that the system of ordinary differential equations assumes that the population is mixing at a higher level than in reality. It is therefore expected that the model will estimate higher levels of immunity. We note, however, that as vaccination coverage increases and becomes dominant in the population, the difference in seroprevalence estimates from our model and (5) should reduce. We also note that (5) does not include seroreversion (related to waning immunity). Inclusion of seroreversion will boost seroprevalence estimates, so the difference between our results would reduce further.

For interest, in Figures 4, 10, and 13 (assuming increases in *κ* values of 20%, no change, and 38%, respectively), we plot the corresponding distribution of protection against the virus of the different types of immunity in the population, over 10-year age groups, including no protection (*S*_1_), some protection 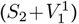, a higher level of protection 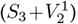, and full protection 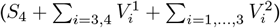.

**Figure 4:**
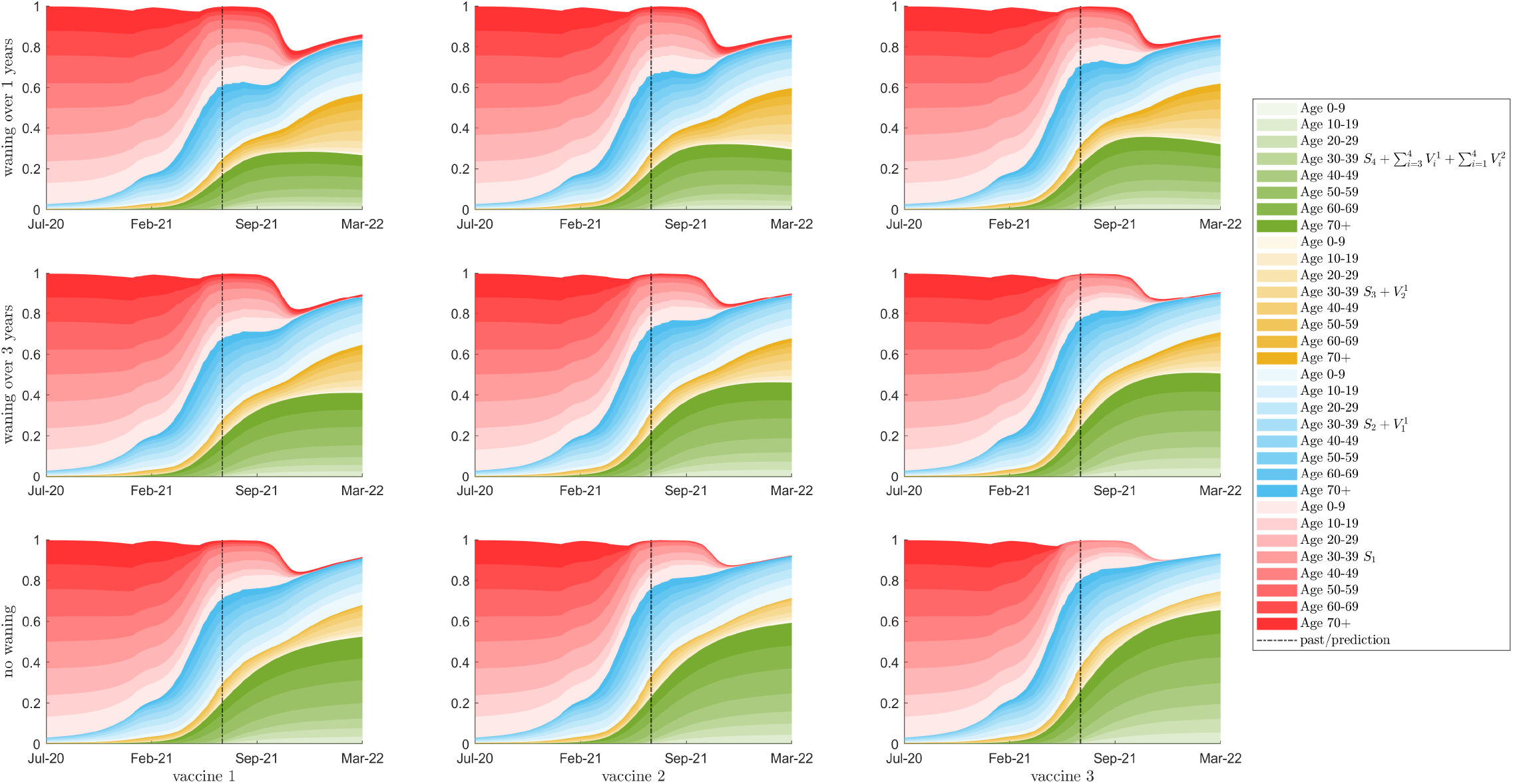
Simulation results with a 20% relaxation of *κ* starting in September 2021 showing immune status of uninfected individuals as a percent of the total population for 10 year age classes are shown with colour intensity corresponding to age class. The colours represent the immunity of the population from green, the fully immune population, through yellow and blue to red, the fully susceptible population. The top row is waning of immunity by one year between consecutive classes; the middle row is waning of immunity of three years; the bottom row is no waning of immunity. Columns left to right represent vaccines 1 to 3, respectively. The vertical bar denotes the beginning of the prediction phase.

In Figures 3-4, 9-10, and 12-13, we observe the effects of waning immunity. Immunity decays from higher immune classes to lower immune classes over time.

### Resurgence

While Figures 2, 8 and 11 show that there is no significant difference between model fits, we do observe divergent daily incidence past September 2021, with the reopening of schools (Figure 8), and when *κ* also increases at this date (Figures 2 and 11). When seroprevalence is high, corresponding to lower rates of waning immunity, we see that resurgence occurs in a milder fashion.

In the top row of Figures 2 and 11, when immunity wanes over 1 year between *V* and *S*, and between consecutive *S* classes (3 years from full protection to full susceptibility), the Fall 2021 resurgence rises to daily incidence levels greater than any counts previously experienced by the Canadian population, no matter the vaccine type (compare columns). Such an occurrence would mean that public health mitigation strategies would need to be implemented in Fall 2021. A booster vaccine program roll-out could also be considered.

When immunity wanes over three years between classes (9 years from full protection to full susceptibility), see Figures 2 and 11, middle row, the resurgence is slightly delayed for all vaccine types, but the resurgence still exceeds any case counts experienced previously. We conclude that, when immunity wanes over nine years (on average) between full protection to full susceptibility, public health mitigation or a booster vaccine campaign will need to be considered. Certainly, for example, if further relaxation in behaviour is experienced, or if there is introduction of new variants of concern, both which increase *κ*.

For interest, we plot the outcomes of our model considering no waning immunity in the bottom row of Figures 2, 8, and 11. Under these assumptions, resurgence is also predicted, but at lower magnitudes. Considering vaccines 2 and 3, we see levels similar to what was previously experienced in Canada’s COVID-19 outbreak.

We note that the resurgence in Fall 2021 is mainly driven by the virus circulation in Summer 2021. Model results show that if relaxation in July 2021 had not occurred, limiting the transmission of the delta VOC, resurgence would be greatly delayed and would have much lower infection levels (results now shown).

Individuals who received the vaccine early in the vaccination program will likely have experienced some effects of waning immunity by September 2021 (see Figures 3-4, 9-10, and 12-13). Resurgence in infections will thus be stronger in these age groups. Figure 5 plots the daily incidence in severe *I*_4_ infections for each vaccine scenario when school is reopened in September and *κ* is increased by 20%. Here, we see that resurgent cases are observed mainly in age groups 50+ when immunity wanes over 1 year between consecutive immune classes (top row). When immunity wanes over 3 years between immune classes, the majority of disease burden is found in age groups 40+. Considering that COVID-19 infection fatality rates increase by age (5), we suggest that a vaccine booster campaign be considered in older age groups.

**Figure 5:**
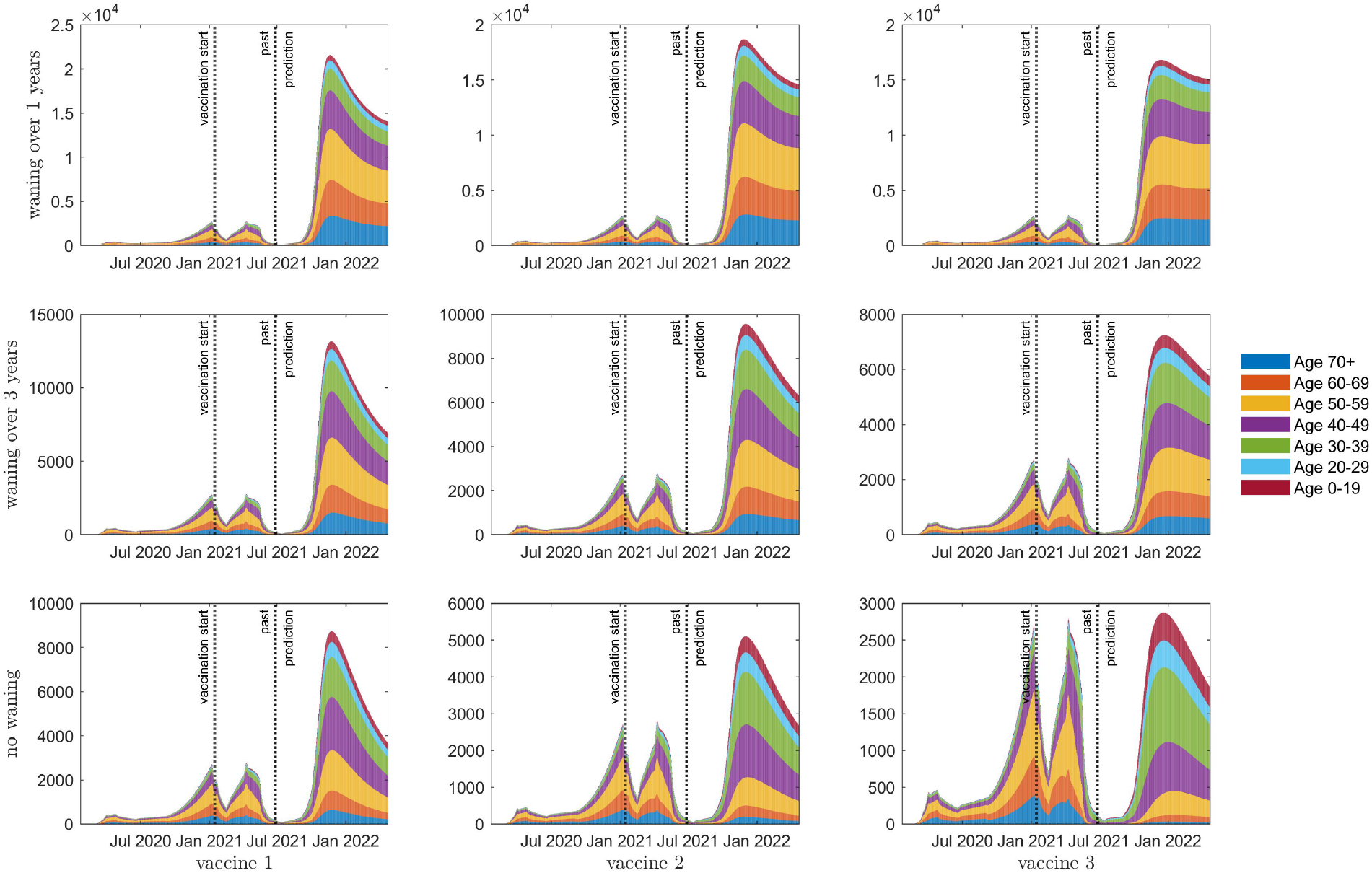
The age structure of daily incidence of severe disease, *I*_4_, over 7 different age groups (see legend and corresponding colours). The top row is waning of immunity by one year between consecutive classes; the middle row is waning of immunity of three years; the bottom row is no waning of immunity. Columns left to right represent vaccines 1 to 3, respectively.

The degree of resurgence is affected by increases in *κ* and school re-opening. Given the higher levels of average daily contacts in younger age groups (see Figure 6, bottom right panel), and given that there is no vaccine yet approved for these ages, we recommend continued use of protective measures in schools, including mask wearing and social distancing. We also recommend that vaccination begin for these ages immediately after a vaccine is approved.

**Figure 6:**
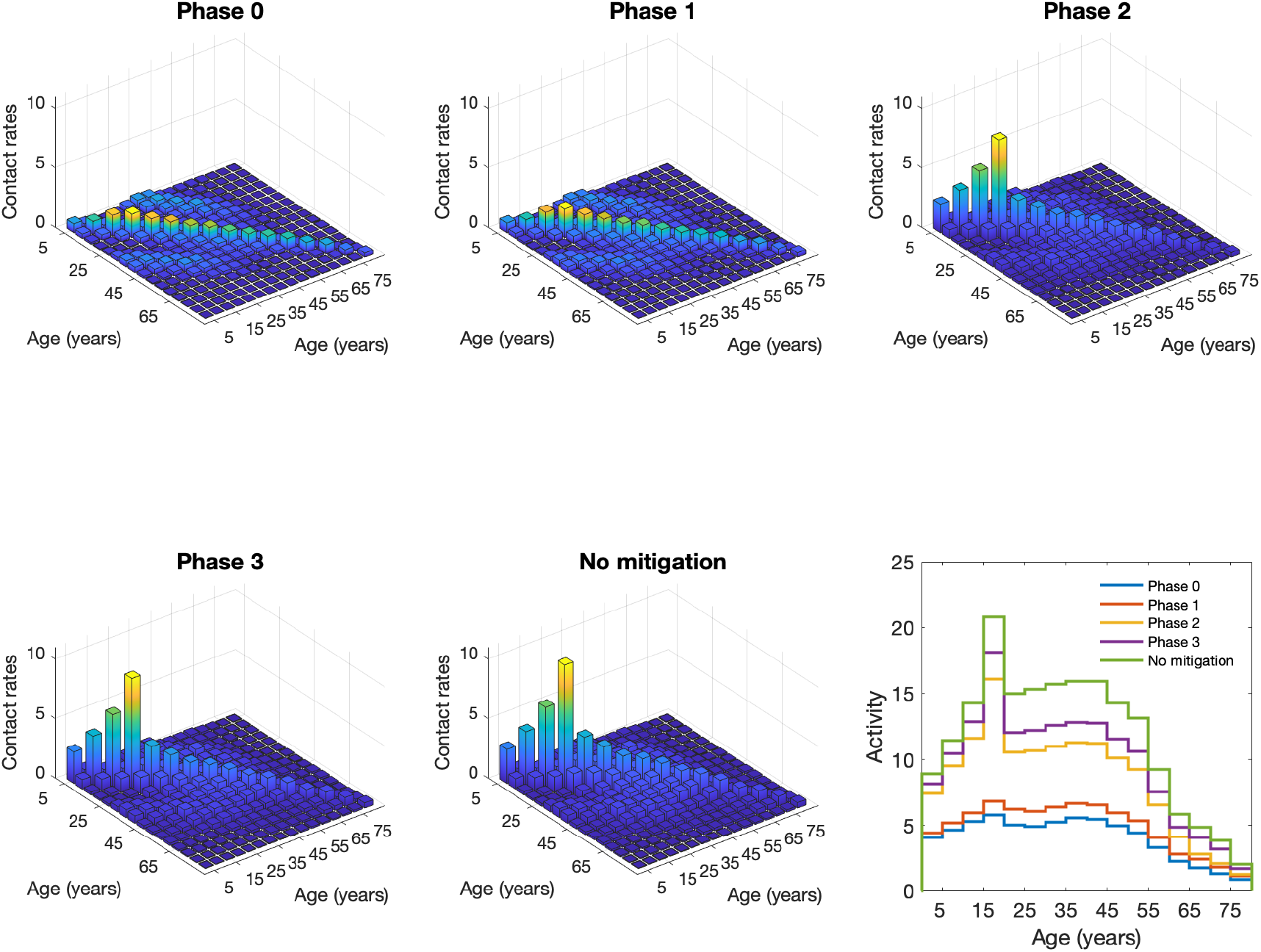
Contact rates and activity under different phases of mitigation. Bar plots show contact rates between age groups under different mitigation strategies, with compliance (*k*) assumed to be 100%. Taller bars, by subplot, have lighter colors. All bar plots have identical y-axis scale for comparison (but are not identical in colour scale). The total activity is shown in the line plot (bottom right).

## Discussion

Our model predicts that 60 to 80% of the Canadian population will have some immunity to SARS-CoV-2 by the end of the vaccination campaign in late Summer 2021. The population is vulnerable to virus resurgence, however, as non-pharmaceutical measures are relaxed allowing spread of the delta VOC. Resurgence degree is augmented by the reopening of school since an approved vaccine for children under 12 is not available in Canada.

The timing and severity of any resurgence is sensitive to the introduction of VOCs, the cessation of personal protective measures and public health mitigation. This sensitivity is highlighted through the comparison of Figures 2 and 11. The possible effects of imported cases are not included and will require further evaluation and, if the virus remains endemic in the population, are unlikely to substantially alter the model outputs according to recent modelling by members of our group (21). As immunity is lost a vaccine booster campaign or the re-introduction of public health mitigation will be required.

## Data Availability

There is no data used in this study.

## Appendix

Perturbations of the contact matrices are implemented to reflect public health mitigation strategies over time. A summary of the perturbations is given in Table 2.

**Table 2:** Contact Perturbation Summary

| Contact mitigation phase description (% reduction in contacts) |  |  |  |
| --- | --- | --- | --- |
| Phase | School Contacts | Other Contacts | Work Contacts |
| 0 | 95% | 90% under 65, 95% over 65 | 75% under 65, 95% over |
| 1 | 95% | 75% | 70% under 65, 95% over |
| 2 | 95% | 40% under 65, 65% over | 70% under 65, 95% over |
| 3 | 15% under 20, 25% over | 40% under 65, 65% over | 35% under 65, 95% over |
| no mitigation | 0% | 0% | 0% |

Table 2: Contact Perturbation Summary
| Contact mitigation phase |  |  |
| --- | --- | --- |
| Start | End | Phase |
| 2020-02-05 | 2020-03-18 | no mitigation |
| 2020-03-18 | 2020-03-31 | 3 |
| 2020-03-31 | 2020-04-25 | 1 |
| 2020-04-25 | 2020-06-19 | 0 |
| 2020-06-19 | 2020-09-01 | 1 |
| 2020-09-01 | 2021-01-08 | 3 |
| 2021-01-08 | 2021-01-22 | 1 |
| 2021-01-22 | 2021-02-19 | 0 |
| 2021-02-19 | 2021-04-09 | 1 |
| 2021-04-09 | 2021-05-14 | 0 |
| 2021-05-14 | 2021-07-15 | 1 |
| 2021-07-15 | 2021-09-01 | 2 |
| 2021-09-01 | 2021-12-31 | 3 |
| 2021-09-01 | 2021-12-31 | 3 |

**Figure 7:**
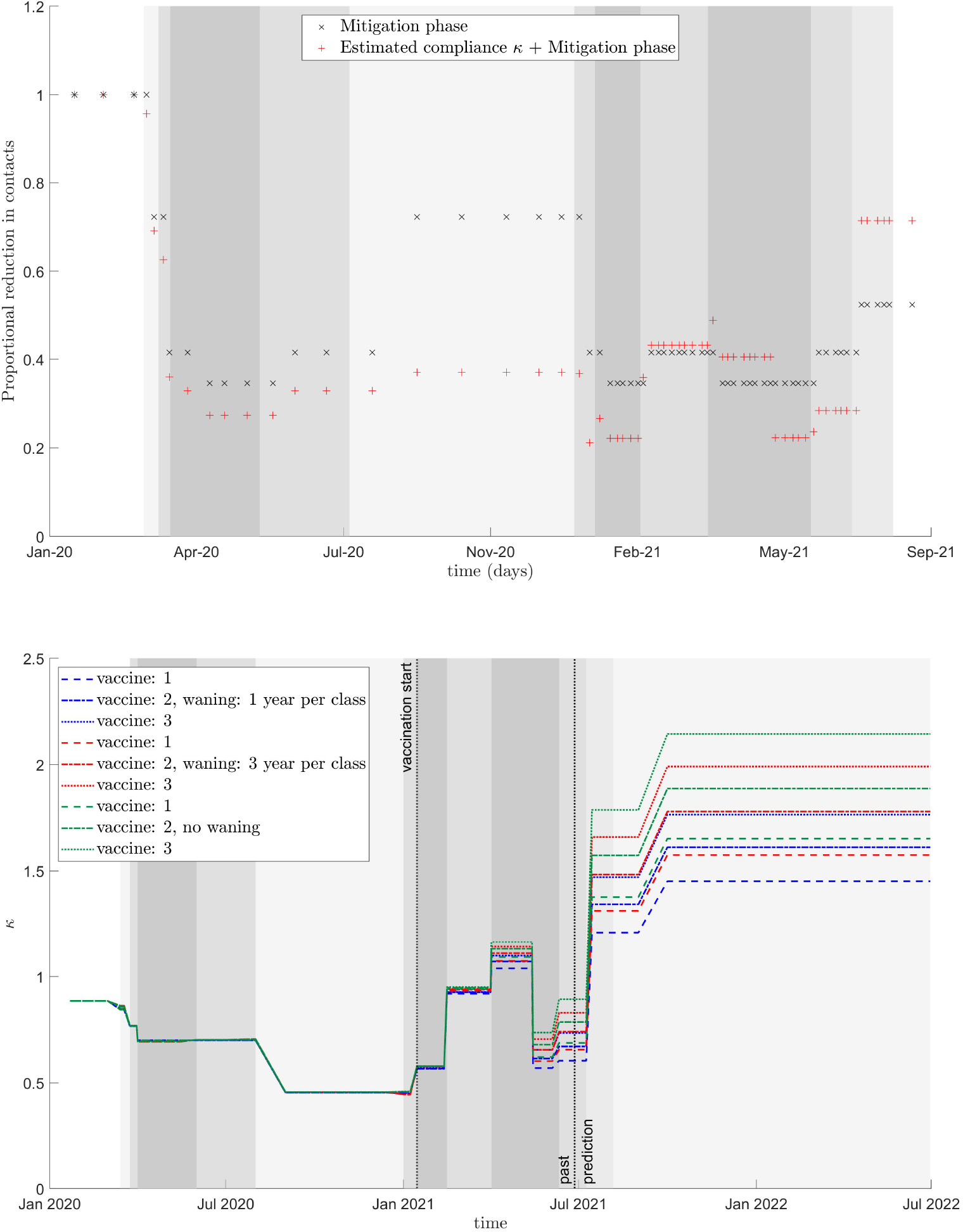
Changes in contact levels over time. (top panel) Average overall contacts compared to no mitigation from modified contact matrices (x’s) plus parmater *κ* (red +’s). (bottom panel) Value of parameter *κ* from the model fit (before black vertical line), and into the future (after black vertical line). A 20% increase in *κ* in September 2021 is assumed here. In both panels, mitigation phases are shown with background shading where darker shading represents more restrictive mitigation, see Table 2 for details.

**Figure 8:**
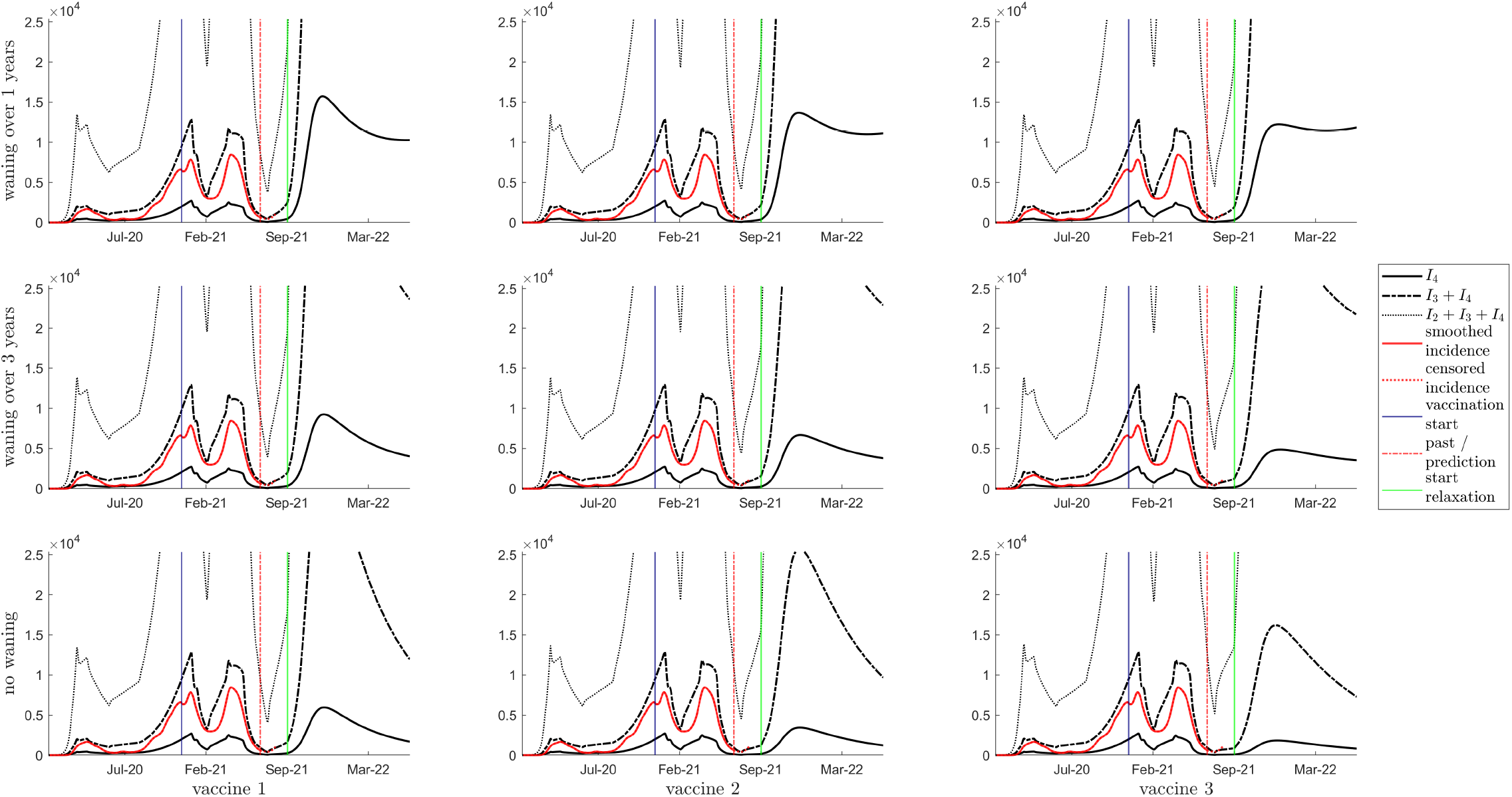
Daily incidence of *I*_4_ (solid line), *I*_3_ + *I*_4_ (dashed line) and *I*_2_ + *I*_3_ + *I*_4_ (dotted line) assuming no relaxation in *κ*, and smoothed incidence data is shown with a solid red line. The vertical blue, red and green lines indicate the start of vaccination, the beginning of the prediction phase, the start of relaxation, respectively. The top row is waning of immunity by one year between consecutive classes; the middle row is waning of immunity of three years; the bottom row is no waning of immunity. Columns left to right represent vaccines 1 to 3, respectively.

**Figure 9:**
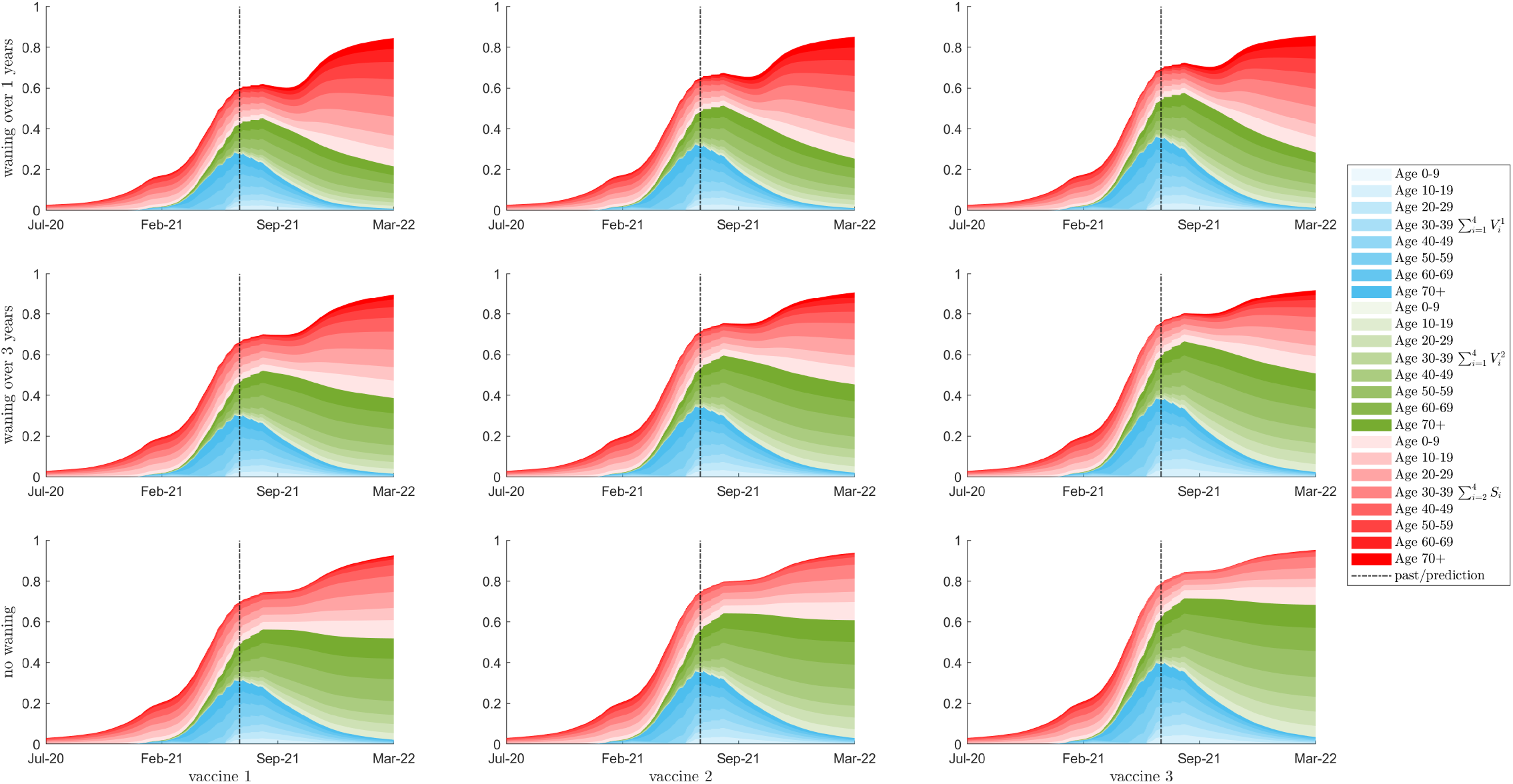
Seroprevalence as a percent of the total population for 10 year age classes are shown with colour intensity corresponding to age class, assuming no relaxation in *κ*. The red region is the sum of a susceptible classes that have been exposed to the virus, either from natural infection or through waning from the vaccinated classes. The blue and green regions show the populations of the first and second dose vaccinated classes respectively. The total population with some immunity (the top of the red region) is equal to the vertical sum of the three blue, red, and green regions. The top row is waning of immunity by one year between consecutive classes; the middle row is waning of immunity of three years; the bottom row is no waning of immunity. Columns left to right represent vaccines 1 to 3, respectively.

**Figure 10:**
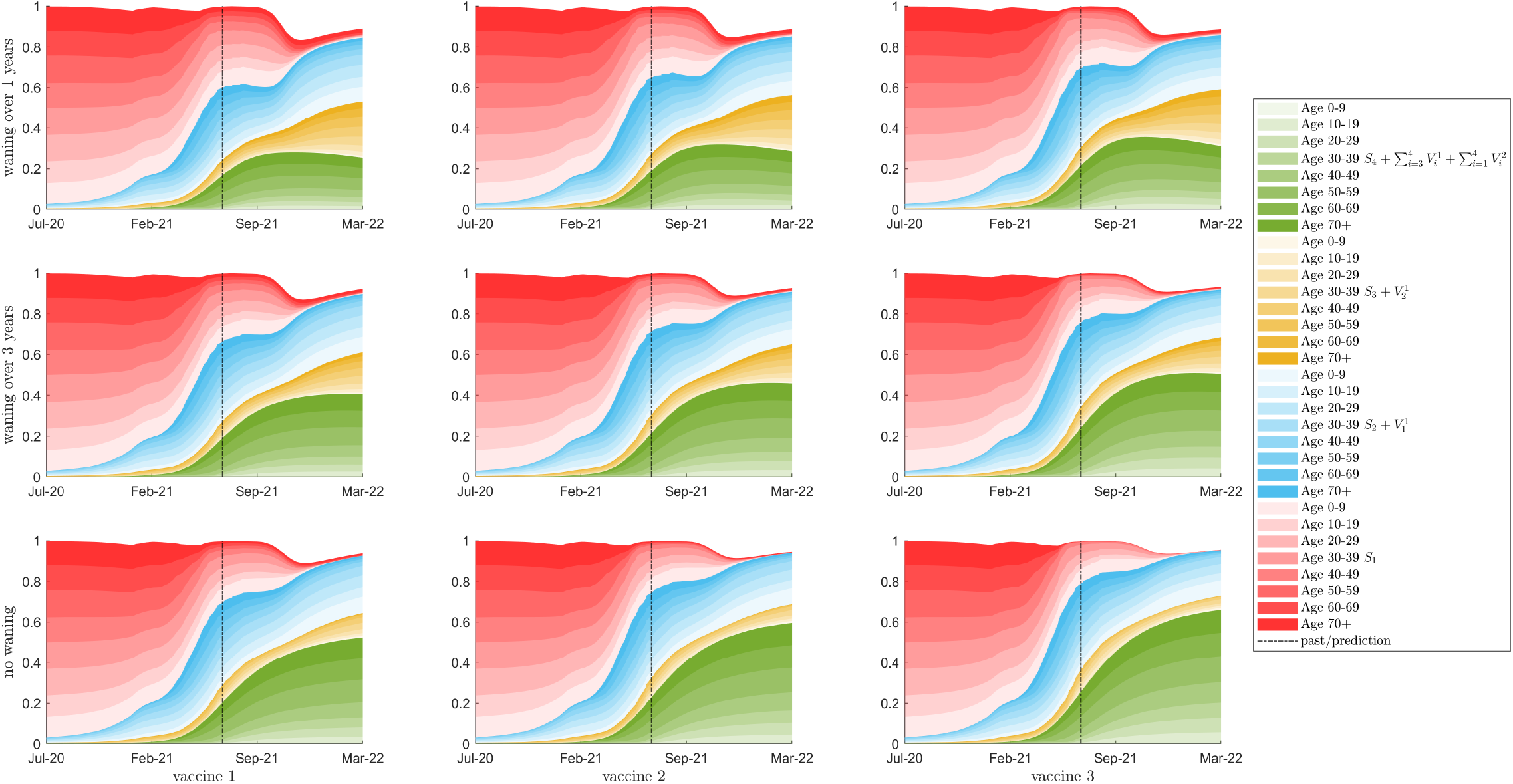
Immune status of uninfected individuals as a percent of the total population for 10 year age classes are shown with colour intensity corresponding to age class, assuming no change in *κ*. The colours represent the immunity of the population from green, the fully immune population, through yellow and blue to red, the fully susceptible population. The top row is waning of immunity by one year between consecutive classes; the middle row is waning of immunity of three years; the bottom row is no waning of immunity. Columns left to right represent vaccines 1 to 3, respectively.

**Figure 11:**
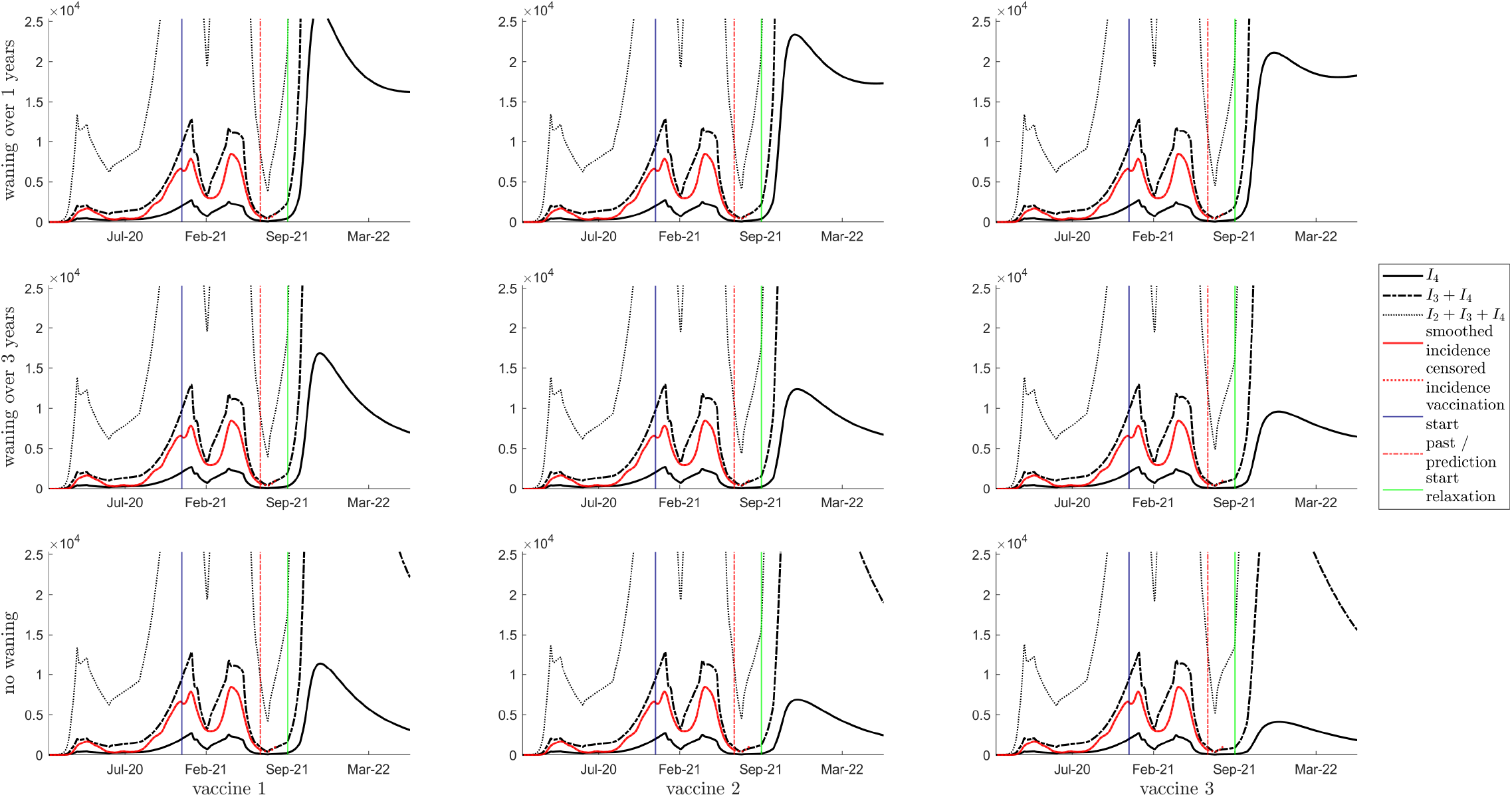
Simulation results with a 38% relaxation of *κ* starting in September 2021 showing daily incidence of *I*_4_ (solid line), *I*_3_ + *I*_4_ (dashed line) and *I*_2_ + *I*_3_ + *I*_4_ (dotted line), and smoothed incidence data is shown with a solid red line. The vertical blue, red and green lines indicate the start of vaccination, the beginning of the prediction phase, the start of relaxation, respectively. The top row is waning of immunity by one year between consecutive classes; the middle row is waning of immunity of three years; the bottom row is no waning of immunity. Columns left to right represent vaccines 1 to 3, respectively.

**Figure 12:**
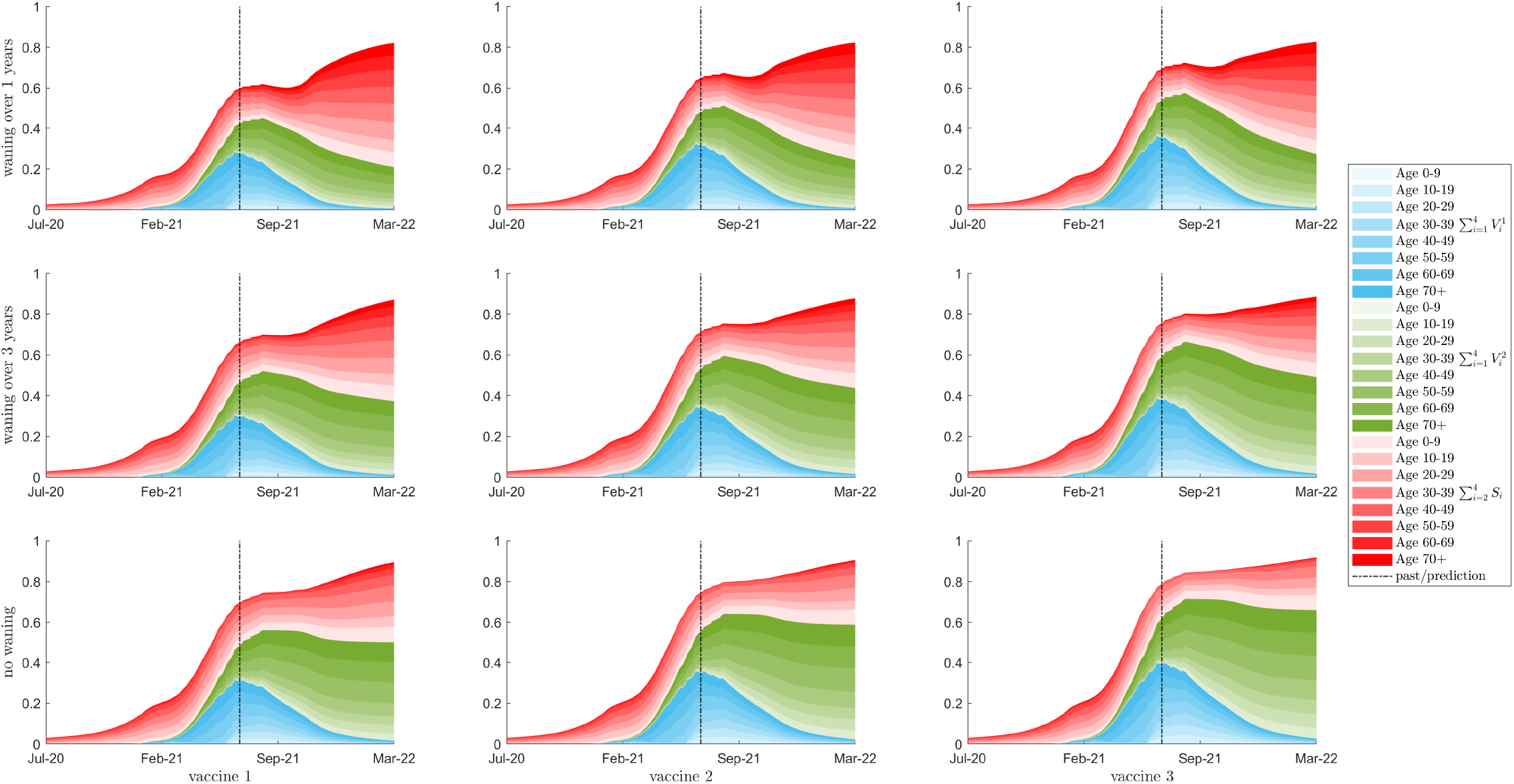
Simulation results with a 38% relaxation of *κ* starting in September 2021 showing seroprevalence as a percent of the total population for 10 year age classes are shown with colour intensity corresponding to age class. The red region is the sum of a susceptible classes that have been exposed to the virus, either from natural infection or through waning from the vaccinated classes. The blue and green regions show the populations of the first and second dose vaccinated classes respectively. The total population with some immunity (the top of the blue region) is equal to the vertical sum of the three blue, red, and green regions. The top row is waning of immunity by one year between consecutive classes; the middle row is waning of immunity of three years; the bottom row is no waning of immunity. Columns left to right represent vaccines 1 to 3, respectively.

**Figure 13:**
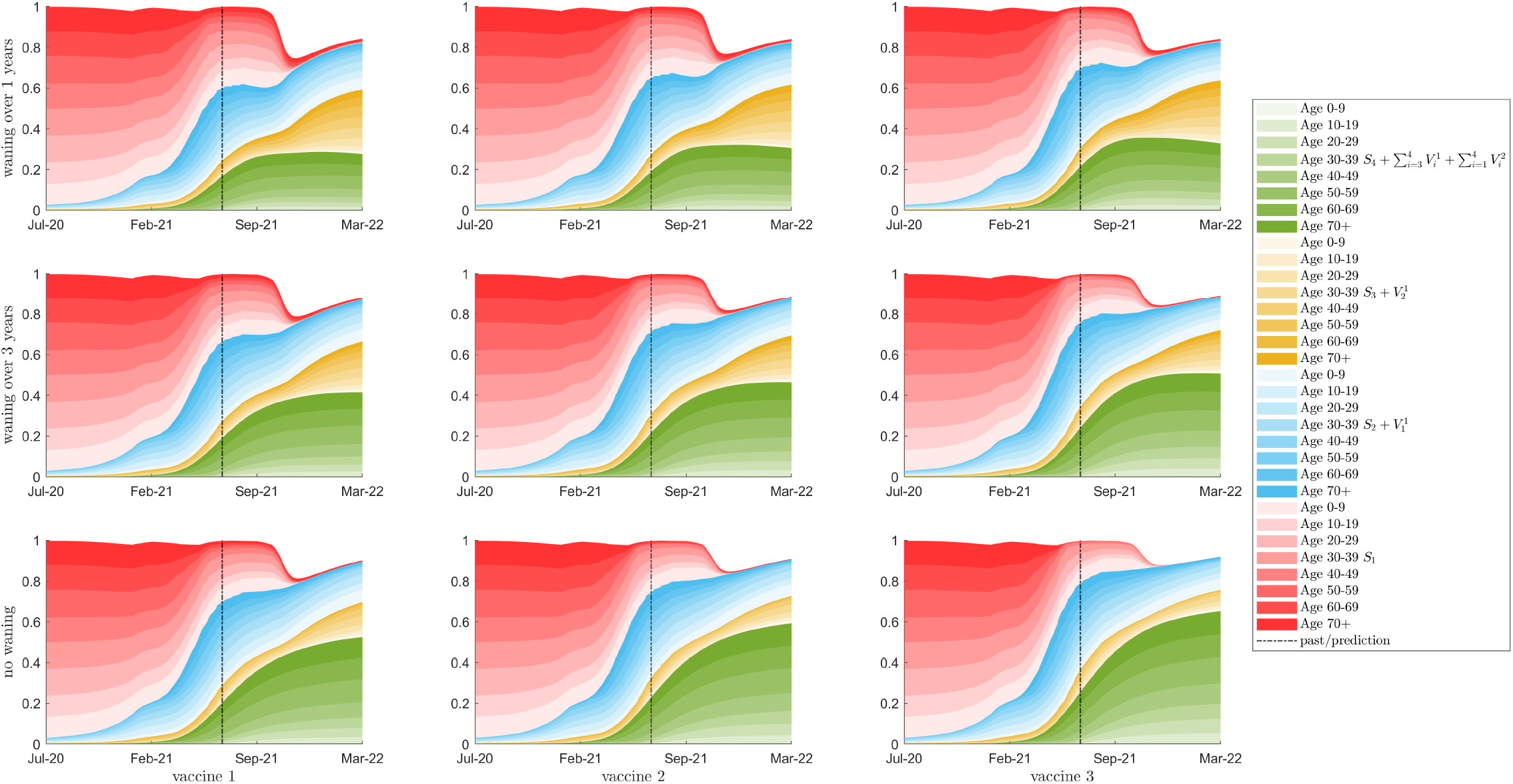
Simulation results with with a 38% relaxation of the k-value starting in September 2021 showing immune status of uninfected individuals as a percent of the total population for 10 year age classes are shown with colour intensity corresponding to age class. The colours represent the immunity of the population from green, the fully immune population, through yellow and blue to red, the fully susceptible population. The top row is waning of immunity by one year between consecutive classes; the middle row is waning of immunity of three years; the bottom row is no waning of immunity. Columns left to right represent vaccines 1 to 3, respectively.

